# Disentangling Symptom Heterogeneity in Large-Scale Psychiatric Text: Domain-Adapted vs. Instruction-Tuned Transformers

**DOI:** 10.64898/2026.02.24.26347006

**Authors:** Giuseppe Varone, Poornima Kumar, Joshua C. Brown, Wadii Boulila

**Author notes:** Corresponding author, (G. Varone). Principal corresponding author. (P. Kumar); (J.C. Brown); (W. Boulila).

## Abstract

Psychiatric disorders are fundamentally challenged by symptom heterogeneity, high comorbidity, and the absence of objective biomarkers, which together result in substantial variability in clinical assessment and treatment selection. Patient-generated language captures rich information about subjective experience and symptom severity, which can be systematically encoded and analyzed using computational models, making it a scalable signal for psychiatric assessment. We compare two approaches: (i) a domain-specialized transformer fine-tuned on clinical language, based on the `Bio-ClinicalBERT` encoder architecture, and (ii) a large-scale instruction-tuned generalist encoder (`Instructor-XL`) used as a frozen feature extractor with a shallow classification head. A corpus of *N* = 151,228 de-identified texts was compiled from five public sources, covering four psychiatric phenotypes: anxiety, depression, schizophrenia, and suicidal intention. Models were evaluated using stratified 10-fold cross-validation with cost-sensitive training, prioritizing imbalance-aware metrics, including Macro-*F*_1_ and Matthews Correlation Coefficient (MCC), over accuracy. `Bio-ClinicalBERT` achieved superior overall performance (Macro-*F*_1_ = 0.78, MCC = 0.6752), indicating more reliable separation of diagnostically overlapping affective categories. In contrast, `Instructor-XL` achieved its highest class-specific performance for schizophrenia (*F*_1_ = 0.798). Explainability analyses suggest that the domain-specialized model places greater weight on clinically relevant terms, whereas the generalist model relies on a broader set of lexical features.

## 1. Introduction

Contemporary psychiatry confronts a fundamental challenge: symptom-based and highly overlapping diagnostic categories often obscure the heterogeneous neurobiological processes underlying mental disorders (1). While clinical evaluation remains heavily dependent on semi-structured clinical interviews using Diagnostic Statistical Manual (DSM) and standardized instruments, such as the Patient Health Questionnaire-9 (PHQ-9) for depression (2), the Generalized Anxiety Disorder-7 (GAD-7) for anxiety (3), and the Positive and Negative Syndrome Scale (PANSS) for psychotic symptomatology—these categorical scores frequently fail to capture the granular, “fuzzy” boundaries of psychiatric distress. Natural Language Processing (NLP) offers a transformative framework for information fusion in mental health by treating human language as a continuous, high-dimensional proxy for mental states (4). By quantifying linguistic patterns, NLP enables “linguistic phenotyping”, which captures the heterogeneity of psychiatric experiences more effectively than traditional scores (5). This paradigm shifts the focus toward inferring latent semantic and affective structures, aligning with generative and Bayesian accounts of cognition that conceptualize mental processes as probabilistic and hierarchical (6). Notably, transformer-based architectures leverage multi-head self-attention mechanisms to encode and integrate multi-scale linguistic features—ranging from local syntactic dependencies to global discourse-level semantics—into a unified high-dimensional vector space. This enables the mapping of unstructured clinical narratives into structured representations that preserve the contextual distinction of psychiatric distress (7; 8). However, the clinical translation of these models is limited by the significant linguistic overlap across disorders. To address this, we propose a framework centered on the fusion of specialized domain knowledge and large-scale semantic priors. We evaluate this framework on an aggregated corpus of *N* = 151,228 samples. Our contributions are threefold:

- **Comparative Representation Fusion:** We contest two paradigms of information extraction: **domain-adaptive fine-tuning** (Bio-ClinicalBERT (9) (10); Model A), which optimizes for specialized clinical likelihoods, and **high-capacity instruction-fine-tuned encoders** (Instructor-XL; Model B) (11), which leverage massive-scale semantic priors. We quantify how these distinct strategies resolve semantic ambiguity in psychiatric discrimination.
- **Bayesian Decision Boundary Optimization:** We employ Bayesian hyperparameter optimization via Tree-structured Parzen Estimators (TPE) to stabilize model calibration. This approach ensures that the fusion of linguistic markers remains robust even under conditions of extreme class imbalance, thereby mitigating the “noise” inherent in naturalistic datasets.
- **Explainable AI (XAI) Verification:** We utilize gradient-based saliency analysis and SHAP (SHapley Additive exPlanations) to interrogate the fused representations. This validation step examines whether model predictions are driven by symptom-related lexical features rather than spurious linguistic patterns. By identifying tokens that make the largest contributions to classification, we assess the extent to which model outputs correspond to established symptom descriptors across diagnostic categories.

We hypothesize that the generalized priors of `Instructor-XL` support more stable identification of sparse phenotypic markers (e.g., schizophrenia), whereas the specialized clinical representations of `Bio-ClinicalBERT` are more responsive to semantic overlap among affective disorders. Furthermore, Bayesian optimization-guided parameter tuning aims to improve the classification across imbalanced and sparsely represented psychiatric categories. The remainder of the manuscript is organized as follows: Section II reviews related work; Section III details the dataset and evaluation matrix; Section IV presents the deep learning architectures and optimization algorithms; Section V presents the performance analysis; Section VI discusses the clinical implications; and Section VII concludes the study.

## 2. Related Work and State of the Art

Over the past decade, research on the application of NLP to psychiatric diagnostics has increased, specifically addressing the distinct information challenges posed by different phenotypes. One primary focus involves identifying sparse linguistic markers in low-frequency or structurally distinct phenotypes, such as schizophrenia (12; 13) and autism spectrum disorders (14), where the challenge lies in detecting deviations in syntactic coherence and thought organization. In contrast, a second research trajectory addresses the high degree of semantic overlap characteristic of affective and internalizing disorders, including depression (15), anxiety (3), and bipolar disorder (16). In these cases, the task shifts from detecting clear structural deviations to resolving “fuzzy” boundary distinctions within a high-dimensional affective feature space. Methodologically, NLP approaches have evolved from traditional morphological and syntactic analyses (17) toward contextualized distributed representations. Although early dictionary-based methods, such as Linguistic Inquiry and Word Count (LIWC), successfully quantified emotional and grammatical features, they are inherently limited by their lexical rigidity. Such models do not account for context-dependent polysemy and discursive variation, including sarcasm, cognitive fatigue, and temporal shifts in affective tone, which are essential for accurate characterization of clinical narratives (18; 19). The introduction of transformer-based architectures marked a paradigm shift toward modeling latent semantic dependencies through self-attention mechanisms. Within this landscape, two competing representational strategies have emerged. Domain-specialized models, such as Bio-ClinicalBERT, prioritize contextual specificity by fine-tuning on biomedical and clinical corpora to enhance sensitivity to subtle psychiatric signals (7). In contrast, high-capacity instruction-tuned LLMs function as static-weight encoders, providing rich semantic embeddings derived from massive-scale generalist priors. However, the extent to which these frozen representations can capture the context-dependent linguistic markers required to express individual symptoms remains an open question (20). Furthermore, as the complexity of the model increases, the hyperparameter surface becomes increasingly non-convex, necessitating robust optimization strategies to ensure model calibration and clinical trust. To address this, current state-of-the-art frameworks integrate Bayesian hyperparameter optimization to stabilize decision boundaries and XAI techniques, such as SHAP, to attribute model variance to specific input tokens (21). To contextualize these advances, Table 1 synthesizes the co-evolution of clinical NLP reviews (Section A) and the underlying architectural formalisms (Section B) that inform current psychiatric linguistic phenotyping. This overview situates our study within the broader landscape of computational psychiatry and information fusion.

**Table 1.** Key reviews in psychiatric NLP and milestones in NLP deep learning architectures.

| Section A: Reviews in Psychiatric NLP |  |  |  |
| --- | --- | --- | --- |
| Reference | Journal / Source | Scope / Focus | Contribution |
| Zhang *et al.* (2022) (22) | *npj Digital Medicine* | NLP for mental illness detection | Trends and methods in psychiatric NLP; emphasizes deep learning and diverse text sources. |
| Le Glaz *et al.* (2021) (23) | *J Med Internet Res* | ML & NLP for mental health | Reviews 58 studies; symptom extraction, severity classification, and clinical integration. |
| Malgaroli *et al.* (2023) (24) | *Translational Psychiatry* | NLP in interventions | Reviews 102 studies; algorithmic/clinical features; NLP×MHI framework. |
| Sheriff *et al.* (2025) (25) | Elsevier systematic review | NLP in youth mental health | Early detection, personalized risk, multimodal approaches. |
| Scherbakov *et al.* (2025) (26) | *JMIR Ment Health* | Depression detection | Methods and ethical considerations in NLP screening. |
| Section B: Milestones in NLP Deep Learning |  |  |  |
| Year / Milestone | Model / Technique | Contribution | Reference / Source |
| 1990s | Rule-based / Statistical NLP | Feature extraction, n-grams, HMMs | Manning & Schütze, 1999 |
| 2013 | Word2Vec | Dense word embeddings, semantic similarity | Mikolov *et al.*, 2013 (27) |
| 2014 | GloVe | Global word co-occurrence embeddings | Pennington *et al.*, 2014 (28) |
| 2015 | RNN / LSTM / GRU | Sequential modeling with memory | Hochreiter & Schmidhuber, 1997; Cho *et al.*, 2014 (29) |
| 2018 | ELMo | Contextualized embeddings (bi-LSTM) | Peters *et al.*, 2018 (30) |
| 2018 | BERT | Transformer-based pretraining, fine-tuning | Devlin *et al.*, 2019 (31) |
| 2019 | GPT-2 | Large autoregressive model | Radford *et al.*, 2019 (32) |
| 2019 | Bio-ClinicalBERT | Domain-adapted clinical pretraining | Alsentzer *et al.*, 2019 (7) |
| 2021 | Instruction-tuned LLMs (Instructor-XL) | Frozen high-capacity encoders | Zhang *et al.*, 2022 (11) |
| 2022–2023 | XAI / Bayesian hyperparameter optimization | Interpretability, clinically meaningful features | Lundberg & Lee, 2017; Ghosh *et al.*, 2025 (33) |

## 3. Materials and Methods

### 3.1. Dataset Description and Source Integration

We developed a unified corpus for psychiatric phenotyping by integrating five heterogeneous data streams, resulting in a dataset across four domains: anxiety, depression, schizophrenia, and suicidal intention. All records reflect linguistic markers of psychiatric phenotypes rather than formal clinical diagnoses, adhering to the principle of “linguistic phenotyping” (5). We implemented a taxonomy harmonization protocol, matching source labels (e.g., Reddit sub-communities and human-annotated stress markers) onto our four target phenotypic domains. A summary of the aggregated sources is provided below:

- **Mental Health Dataset:** A corpus of nearly 300,000 records linking linguistic patterns and sentiment scores (Automated Readability Index, Flesch-Kincaid) to psychiatric and behavioral indicators, including economic stress, isolation, and treatment history (https://www.kaggle.com/datasets/bhavikjikadara/mental-health-dataset/data; CC BY 4.0).
- **Dreaddit Corpus (Supervised Stress Analysis):** A corpus of 190,000 posts from five Reddit domains (abuse, anxiety, financial, PTSD, social) with 3,500 human-annotated segments providing binary stress labels (1 = stress, 0 = no stress) (34) (Apache 2.0).
- **SuicideWatch Archive (Longitudinal Risk Detection):** A dataset of 232,074 posts (2008–2021) designed to detect transitions between suicidal intention and neutral states, using a balanced distribution of posts (116,037 suicidal vs. 116,037 non-suicidal) from the r/SuicideWatch and r/teenagers subreddits (https://www.kaggle.com/datasets/nikhileswarkomati/suicide-watch).
- **Reddit Cleaned Dataset (Taxonomy Harmonization):** Approximately 7,650 posts from depression-related subreddits, with a roughly balanced distribution between mental health indicators and neutral controls, harmonizing categorical taxonomies across sources (https://www.kaggle.com/code/ahmedalbaz031/the-depression-dataset-analysis; CC BY-SA 4.0).
- **Pandemic-Era Collection (RMHD):** Nearly 1 million posts (2018–2020, updated to 2022) from 28 mental health subreddits (e.g., r/Anxiety, r/Depression, r/BipolarReddit), providing longitudinal coverage to examine temporal dynamics of psychiatric language (https://www.kaggle.com/datasets/f995248f9f3ed770655c1d2f73d38bb6555ea2fcbb1055f50f5354e3c6c4100d).

Lexical density was quantified (mean ± SEM), revealing systematic differences in narrative verbosity across phenotypes (Figure 5a). To characterize the latent representational geometry of the corpus, document embeddings were projected into a two-dimensional space using Uniform Manifold Approximation and Projection (UMAP) (Figure 5b). This analysis revealed that while psychiatric categories occupy distinct semantic clusters, there is significant transdiagnostic overlap, particularly between anxiety and depression. Finally, a unigram/bigram frequency analysis was conducted to extract the lexical profiles characterizing each domain, providing a grounded reference for the downstream transformer-based modeling (Figure 5c).

### 3.2 Information Fusion and Mathematical Formalization

We formalize multi-source psychiatric text fusion as learning a single predictive mapping from heterogeneous natural-language samples to a unified phenotypic label space. Let the aggregated corpus be

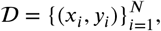

where *x*_*i*_ is the raw text and *y*_*i*_ ∈ 𝒴 = {0, …, *K* − 1} is the harmonized psychiatric label with *K* = 4 (anxiety, depression, schizophrenia, and suicidal intention). Each sample is additionally associated with a source indicator *s*_*i*_ ∈ 𝒮 (e.g., a specific subreddit or component dataset), where 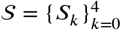.

#### Label harmonization as a source-conditional mapping

Each source *s* provides native labels 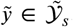. We define a deterministic harmonization map 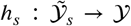 and construct the unified label 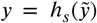. This induces an effective label-noise channel when source taxonomies only partially align; a convenient abstraction is 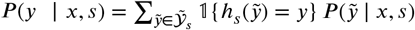.

##### Multi-source fusion as a mixture distribution

We model the fused corpus as draws from a finite mixture of source-conditional joint distributions:

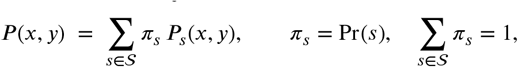

where *P*_*s*_(*x, y*) = *P* (*x, y* ∣ *s*) denotes the source-conditional data-generating distribution and *π*_*s*_ reflects the empirical prevalence of source *s* in the aggregated dataset (typically *π*_*s*_ ≈ *N*_*s*_/*N*). This mixture formalization describes the empirical aggregation of heterogeneous sources and does not assume identical sampling mechanisms across domains. Accordingly, training optimizes performance under the empirical mixture induced by {*π*_*s*_}, enabling a single predictive model to operate over heterogeneous psychiatric text sources.

#### Mixture-risk decomposition and source contributions

Under the mixture model, the population risk of a classifier *f*_*θ*_ under a loss *ℓ* decomposes as a convex combination of source-wise risks:

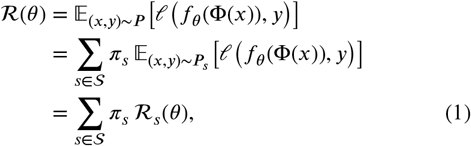

where the source-wise risk is

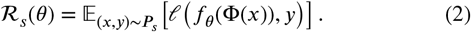

This decomposition shows that optimization is biased toward high-prevalence sources (large *π*_*s*_), which is critical when sources differ in annotation protocol, lexical style, or symptom expression. It also provides a principled lens for contrasting specialist adaptation (Model A) and generalist priors (Model B): Model A primarily minimizes per-domain risk, ℛ_*s*_(*θ*), through source-adaptive representations that capture lexical and clinical regularities. Model B, however, reduces estimator variance by incorporating global semantic priors, leading to more stable estimates of 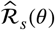 under low-resource conditions.

#### Cross-source generalization under domain shift

To formalize why performance may differ across heterogeneous sources, consider a *target* source *t* ∈ 𝒮 (e.g., a held-out dataset) and a *training mixture distribution*

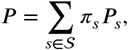

defined as a weighted combination of source distributions {*P*_*s*_}_*s*∈𝒮_ with mixture weights {*π*_*s*_}_*s*∈𝒮_ . For a hypothesis class ℋ, a standard domain adaptation bound characterizes the role of representation quality under distribution shift:

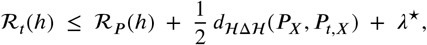

where *d*_ℋΔℋ_ (·, ·) denotes the ℋΔℋ-divergence between input marginals *P*_*X*_ and *P*_*t,X*_, and

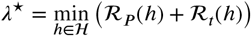

captures the minimal joint risk when labeling functions differ across domains. This bound provides an interpretation of how domain-adaptive fine-tuning (Model A) may decrease ℛ_*P*_( *h*) by specializing to clinical semantics, while instruction-tuned embeddings (Model B) may reduce effective discrepancy *d*_ℋΔℋ_ by providing broader, more stable semantic features across diverse linguistic styles.

#### Stratified evaluation protocol

To improve the stability of performance estimates under class imbalance, we implement a stratified partitioning protocol:

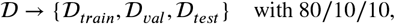

such that the empirical label distribution is preserved across subsets:

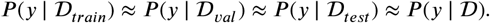

Specifically, 80% of the corpus is allocated to the training set 𝒟train, and the remaining 20% is split evenly between a validation set 𝒟val (used for hyperparameter tuning) and a held-out test set 𝒟_test_ (used for final evaluation), resulting in a 80/10/10 partition.

##### Cost-sensitive learning under class imbalance

Given the highly imbalanced empirical class distribution *P* (*y*), we employ a cost-sensitive loss. Let *n*_*j*_ be the number of training samples in class *j* and *N*_*train*_ = | 𝒟_*train*_|. We compute normalized importance weights

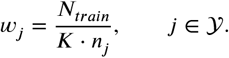

The model *f*_*θ*_ is optimized to minimize the weighted empirical risk:

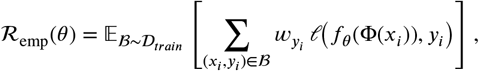

where Φ(·) denotes the transformation from raw text to model input (tokenization and embedding) and *ℓ*(·, ·) is the sparse categorical cross-entropy. Concretely, for a mini-batch ℬ ⊂ 𝒟 _*train*_, the loss is

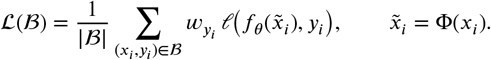

Note that ℛ _emp_(*θ*) ≈ 𝔼_ℬ_ [ℒ ( ℬ)] under mini-batch sampling. All preprocessing, stratified splitting, and class-weight computations were implemented in Python using `scikit-learn` (v1.3.0). Figure 2 illustrates the end-to-end modeling pipeline, while Table 2 summarizes the distribution of psychiatric categories across partitions.

**Table 2.** Distribution of Psychiatric Categories Across Experimental Dataset Partitions.

| Partition | Total (*n*) | Anxiety | Depression | Schizo. | Suicidal |
| --- | --- | --- | --- | --- | --- |
| Training (80%) | 120,982 | 4,580 | 13,345 | 800 | 102,257 |
| Validation (10%) | 15,123 | 572 | 1,668 | 100 | 12,783 |
| Testing (10%) | 15,123 | 573 | 1,668 | 100 | 12,782 |
| **Full Dataset** | **151,228** | **5,725** | **16,681** | **1,000** | **127,822** |

**Figure 1:**
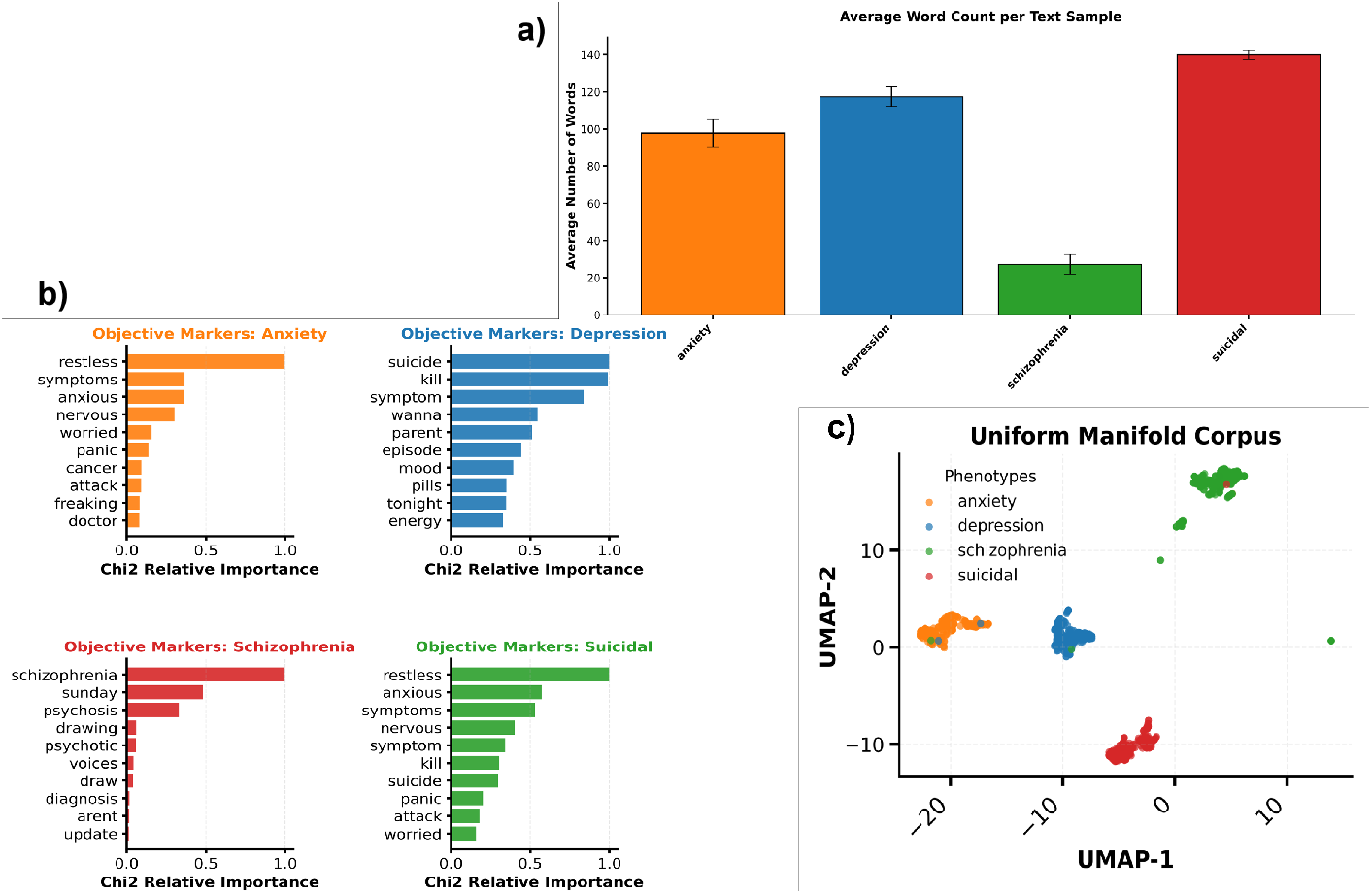
Linguistic and geometric structure of the psychiatric text dataset. The top panel shows the average document length (mean ± SEM) across psychiatric categories, revealing systematic differences in narrative verbosity between anxiety, depression, schizophrenia, and suicidality. Bottom-left panels depict the most frequent symptom-related terms within each category, highlighting phenotype-specific lexical profiles (e.g., anxiety-related arousal terms, depressive affective language, psychosis-related perceptual disturbances, and suicide-related intention). The bottom-right panel shows a UMAP projection, a semantic continuum, not clusters of how clinical notes encode overlapping symptoms.

**Figure 2:**
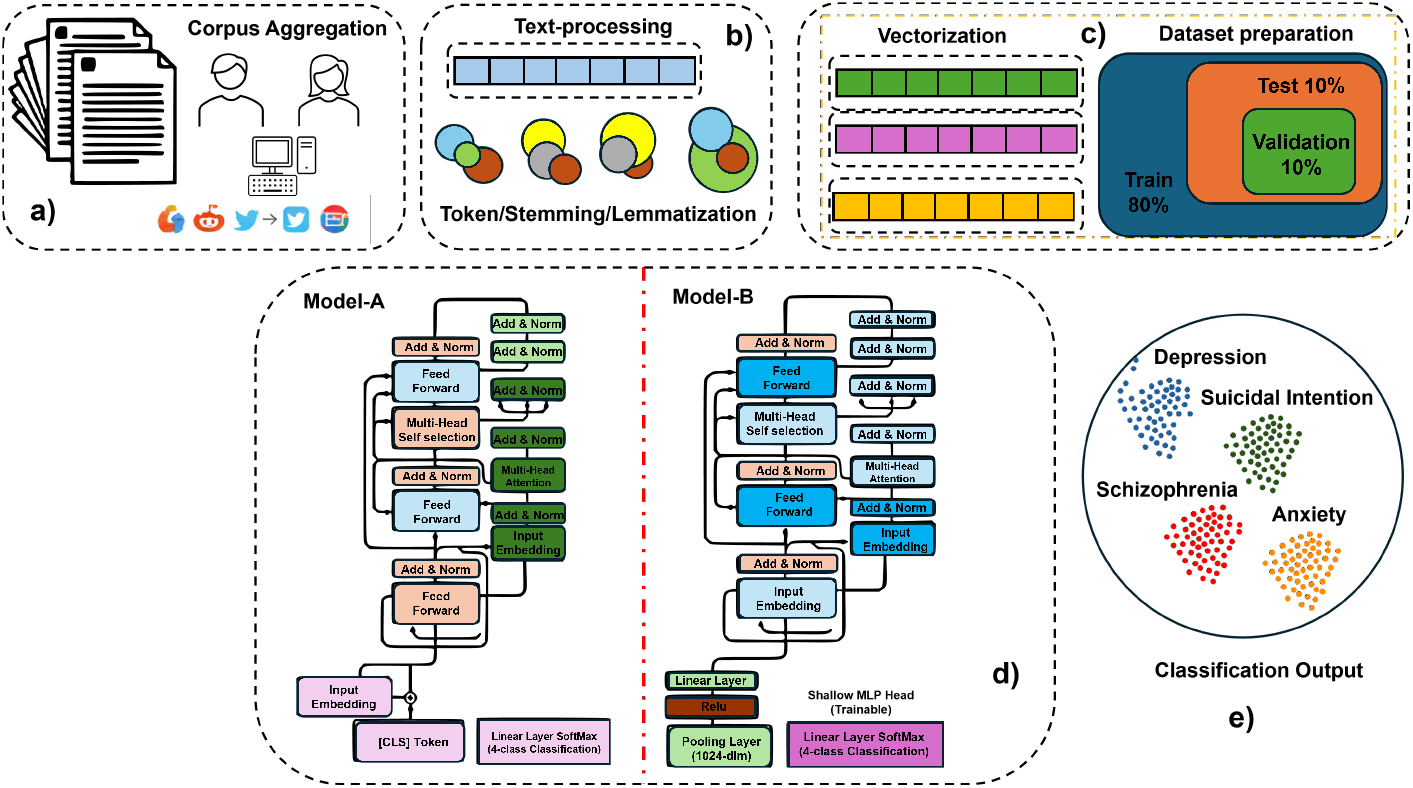
Illustrative example of the end-to-end transformer-based psychiatric text classification framework. Raw psychiatric text is tokenized and mapped to embeddings, optionally augmented with a special token. Two encoder variants are shown: a specialist Model (left) that performs direct classification and a generalist Model (right) that learns shared representations across phenotypes. The data are split into training (80%), test (10%), and validation (10%). Latent embeddings form separable clusters corresponding to anxiety, depression, schizophrenia, and suicidal phenotypes.

### 3.3. Evaluation Metrics

Model performance was evaluated at both global and class-specific levels. Overall classification accuracy on the held-out test set served as a primary metric:

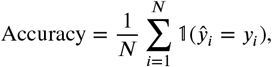

where 𝟙(·) equals 1 if the predicted label *ŷ*_*i*_ matches the true label *y*_*i*_, and 0 otherwise. Accuracy provides an overall measure of correctness but can be misleading when classes are imbalanced. To account for class-specific performance, we computed precision (*P*_*c*_), recall (*R*_*c*_), and F1-score (*F* 1_*c*_) for each class *c* ∈ {0, 1, …, *C* − 1}:

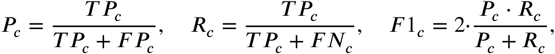

where *T P*_*c*_, *F P*_*c*_, and *F N*_*c*_ denote the number of true positives, false positives, and false negatives for class *c*, respectively. Macro-averaged metrics were computed to summarize performance across all classes:

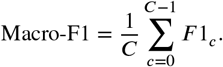

Confusion matrices (*CM* ∈ ℝ^*C*×*C*^ ) were computed to visualize inter-class misclassification patterns:

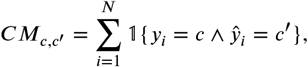

where 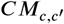 counts the number of instances with true label *c* predicted as *c*^′^. These metrics informed the selection and evaluation of the model, particularly for minority classes.

## 4. Deep Learning

### 4.1. Proposed Architecture: Bio-ClinicalBERT (Model A)

We implemented a Bio-ClinicalBERT model to classify psychiatric texts. Bio-ClinicalBERT is a domain-adapted transformer encoder trained in clinical narratives from the MIMIC-III corpus, providing robust embeddings of medical and psychiatric terminology. Figure 3 illustrates the architecture of Model A.

**Figure 3:**
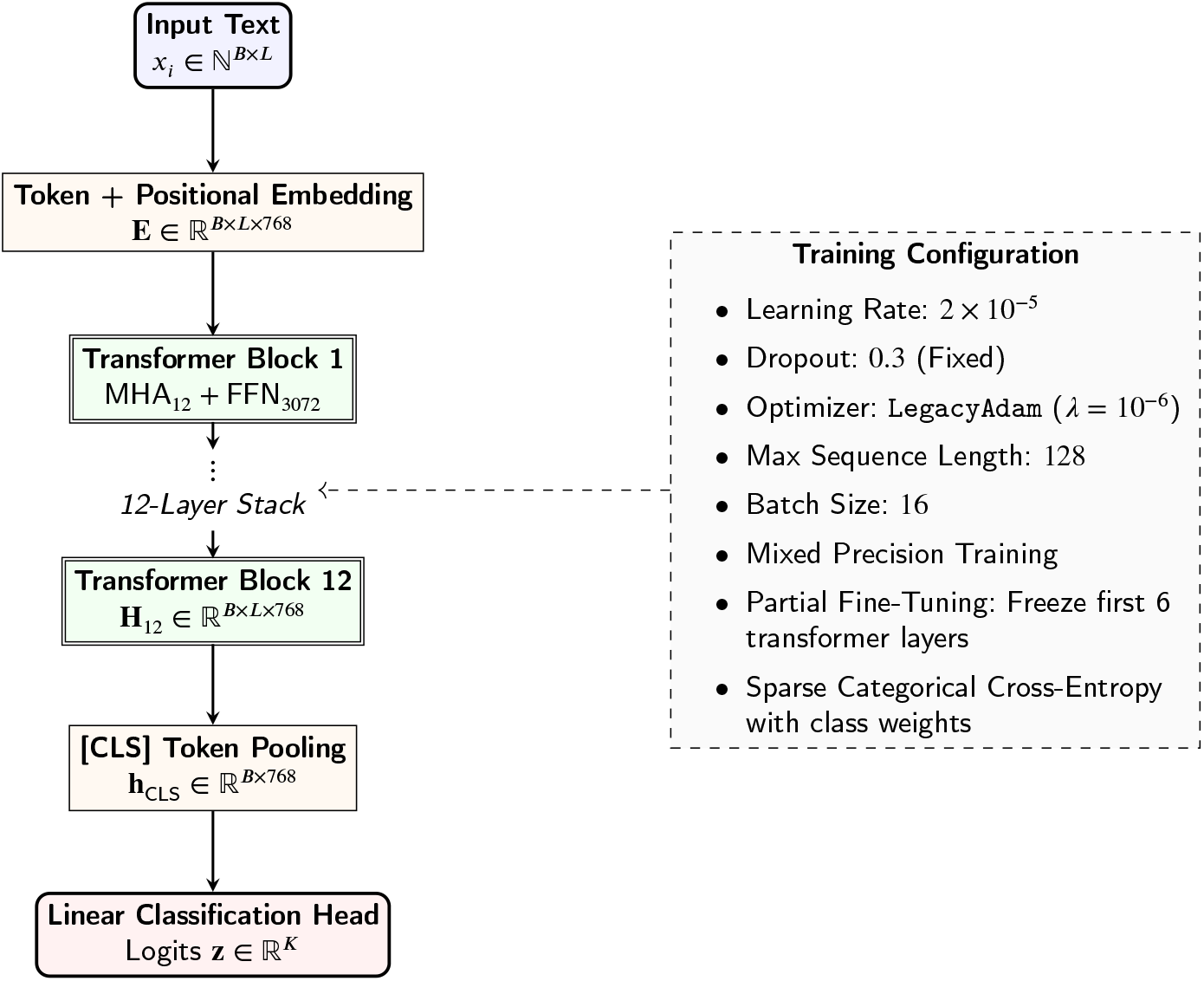
Model A: Partially fine-tuned Bio-ClinicalBERT pipeline. The lower six transformer layers are frozen; the upper layers and classification head are optimized for psychiatric text classification.

**Figure 4:**
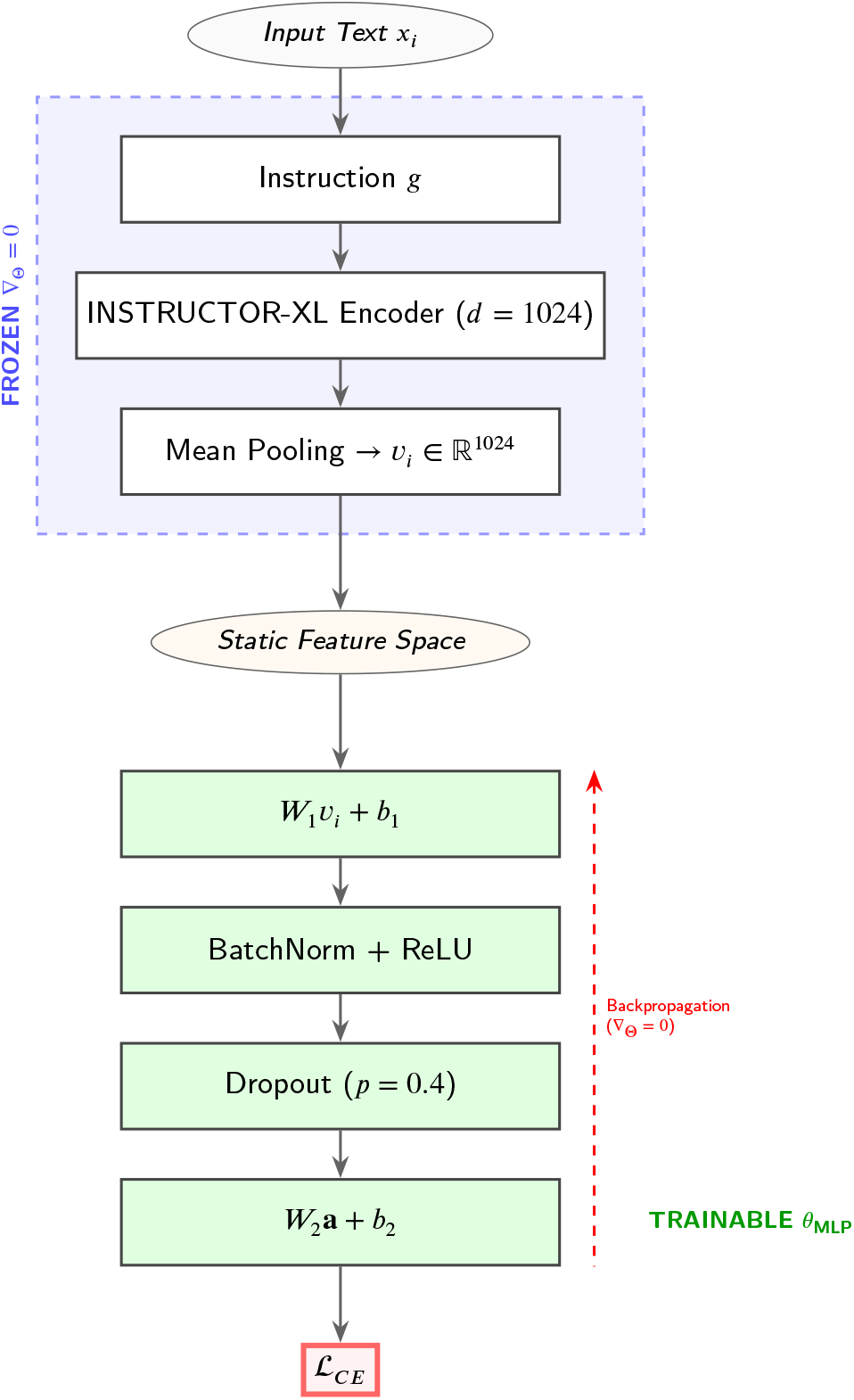
Model B architecture. Input text is encoded using a frozen INSTRUCTOR-XL encoder to produce a 1024-dimensional static representation. Only the MLP classification head is trained using cross-entropy loss, while encoder parameters remain fixed.

**Figure 5:**
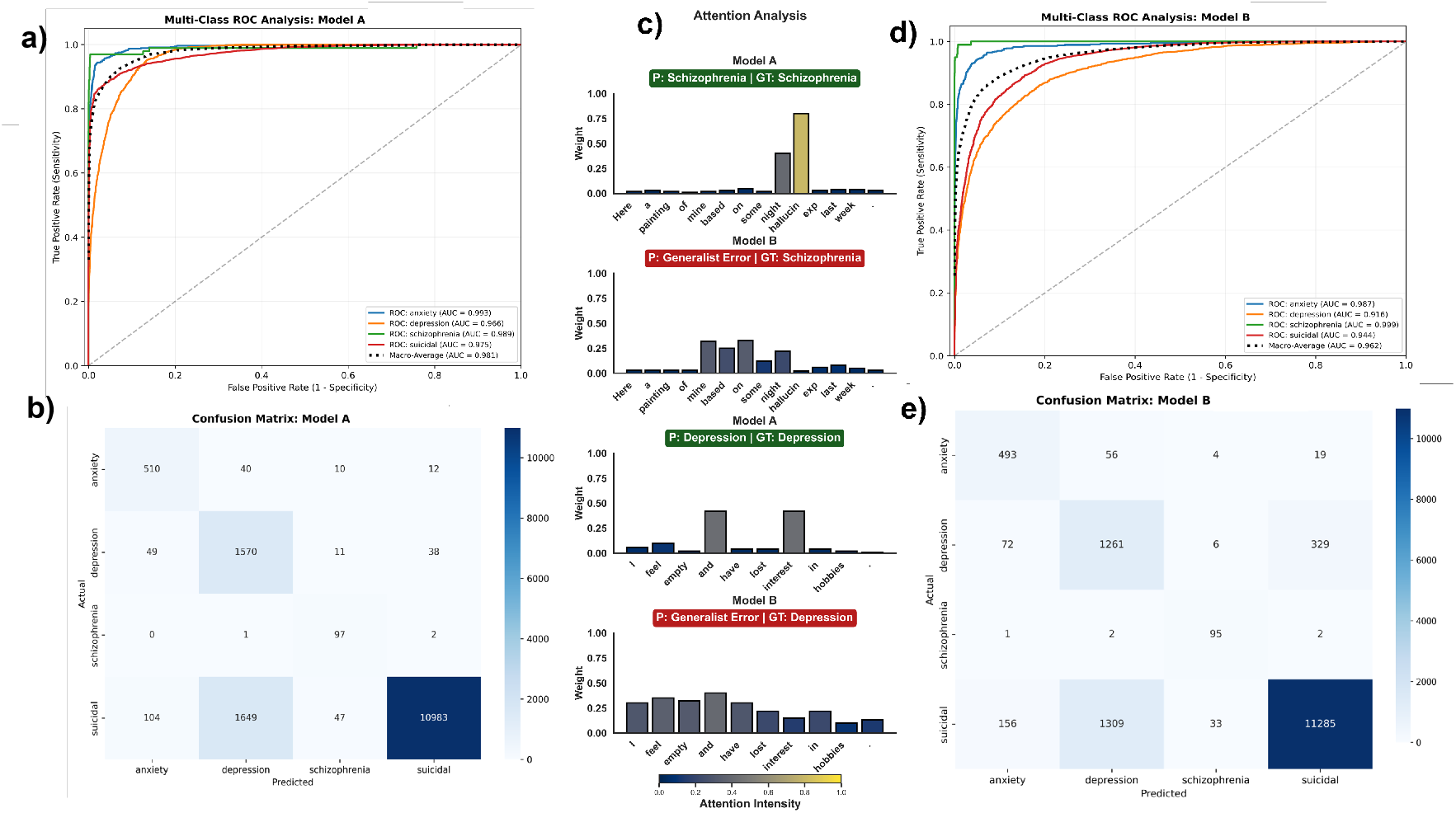
Comparative performance and mechanistic interpretability of Specialist (Model A) and Generalist (Model B) classifiers. Top panels show multi-class ROC curves for anxiety, depression, schizophrenia, and suicidality, including macro-averaged performance, highlighting superior class-specific discrimination in Model A relative to Model B. Bottom panels illustrate the corresponding confusion matrices, suggesting reduced cross-psychiatric misclassification for the Specialist model, particularly for depression and suicidality. Central panels present representative self-attention weight distributions for correctly classified and failure cases, demonstrating that Model A exhibits symptom-specific token weighting, whereas Model B shows more diffuse attention patterns associated with boundary failures. Together, these results indicate that specialized modeling yields improved psychiatric class separability and more interpretable symptom-level representations compared with a generalist approach.

The input corpus consists of variable-length sequences *l* ≤ *L*, where *L* = 128 denotes the maximum sequence length. Each sequence is tokenized and mapped to discrete token indices,

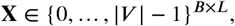

where *B* is the batch size and |*V* | is the vocabulary size. Sequences shorter than *L* are post-padded with a `[PAD]` token to ensure uniform length across the batch.

To prevent self-attention from attending to padding positions, a binary attention mask

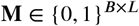

is defined, where **M**_*V,l*_ = 1 indicates a valid token and **M**_*V,l*_ = 0 indicates padding. This mask is converted to an additive attention mask

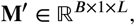

where padding positions are assigned a large negative value (e.g., −∞) and broadcast across query positions. The masked self-attention operation is given by

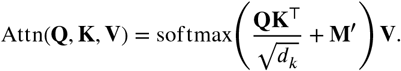

Token embeddings are summed with positional encodings to produce the input representation

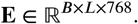

These embeddings are processed through a stack of 12 Transformer blocks. The bottom six blocks are frozen during training to preserve pretrained linguistic representations, while the top six blocks are fine-tuned to adapt the model to the psychiatric text domain. The final hidden representations are denoted by

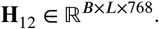

A sequence-level representation is obtained by extracting the hidden state corresponding to the [CLS] token (located at the first sequence position),

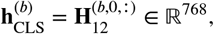

where superscript (*b*) indexes the *b*-th batch element. These representations are passed to a linear classification head, producing logits

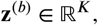

where *K* denotes the number of psychiatric outcome classes.

#### 4.1.1 Training Procedure

In Table 3, we summarize the training configuration hyperparameters for Model A.

**Table 3.** Training Configuration and Hyperparameters.

| Component | Specification / Value |
| --- | --- |
| Loss Function | Sparse Categorical Cross-Entropy (weighted for class imbalance) |
| Optimizer | LegacyAdam with weight decay $\lambda = 10^{-6}$ |
| Batch Size | 16 |
| Learning Rate | $2 \times 10^{-5}$ |
| Dropout | 0.3 |
| Mixed Precision Training | Enabled (GPU speedup) |
| Callbacks | EarlyStopping (patience 2, restore best weights)<br>ReduceLROnPlateau (factor 0.2, patience 1, min LR $10^{-7}$) |

The partial fine-tuning strategy balances computational efficiency and domain adaptation by freezing lower layers to retain clinical pretraining knowledge while licensing higher layers to learn psychiatric-specific linguistic patterns from online text.

#### 4.1.2. Transfer Learning and Domain Adaptation

We use transfer learning to map general-purpose linguistic features into a psychiatric-specific latent space. General models (BERT, RoBERTa) (35; 10) capture broad syntax but lack the priors to resolve clinical semantic ambiguities. Bio-ClinicalBERT (36; 7), pre-trained on electronic health records (EHR) and initialized from BioBERT, produces contextual embeddings optimized for psychiatric terminology. Its fine-tuned attention mechanisms generate high-dimensional vectors that capture embedded meaningful features while minimizing bias from general-language models, providing a representations for integration with neurophysiological measures.

#### 4.1.3. Stochastic Optimization and Hyperparameter Tuning

To optimize the fusion of clinical priors and task-specific semantics, we employed Bayesian optimization via the Tree-structured Parzen Estimator (TPE) (37). The search space Λ was defined in the learning rate *η* ∈ [10^−6^, 10^−4^], the dropout rate *δ* ∈ [0.1, 0.5], and the batch size *B* ∈ {8, 16, 32}. We used a weighted sparse categorical cross-entropy loss to mitigate the effects of class imbalance in the psychiatric corpus.

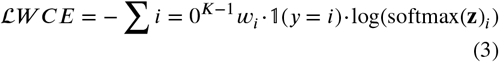

where *w*_*i*_ denotes the normalized inverse frequency of the class *i*.

##### Algorithm 1 Bayesian Hyperparameter Optimization with TPE

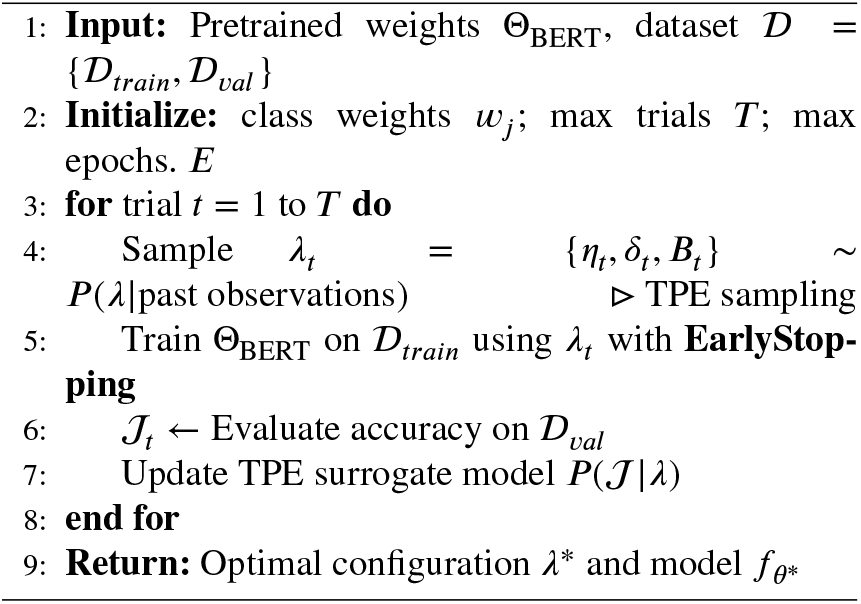

#### 4.1.4. Explainable AI (XAI) and Interpretability

To interpret the decision-making process of the model, we employ KernelSHAP to approximate the Shapley values *ϕ*_*i*_, quantifying the contribution of each linguistic feature to the final diagnosis prediction. Given the high dimensionality of the transformer latent space, we estimate the value function *v*(*S*) using interventional expectations:

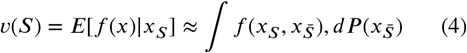

By replacing missing features with samples from a back-ground distribution 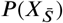, we achieve a tractable estimation of feature importance. This allows us to map the abstract embeddings back to specific clinical tokens, ensuring the model’s outputs are aligned with established psychiatric semiology. The SHAP quantifies feature importance via Shapley values:

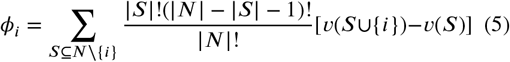

where *v*(*S*) is the prediction model for the characteristic coalition *S*. For high-dimensional embeddings, we use KernelSHAP with interventional expectation:

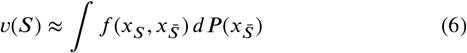

- **Interventional Expectation:** Replace missing features with samples from a background distribution 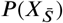 to estimate Shapley values.

KernelSHAP approximates Shapley values by fitting a weighted linear model to sampled feature coalitions, where the contribution of each coalition is computed via the interventional expectation. This approach allows tractable estimation of feature importances in high-dimensional feature spaces, such as the embeddings derived from our large-scale psychiatric text corpus.

### 4.2. Alternative Architecture: Frozen Instruction-Tuned Encoder + MLP Classifier (Model B)

To contrast the efficacy of static high-dimensional embeddings against fine-tuned representations, we implemented Model B: a frozen instruction-tuned transformer encoder coupled with a Multi-Layer Perceptron (MLP). We utilized INSTRUCTOR-XL (11), a 1.5-billion-parameter encoder. Each input *x*_*i*_ is prepended with a domain-specific instruction *g*: *“Represent the clinical statement for psychiatric classification”*. The encoding process is defined as:

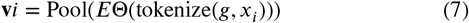

where *E*_Θ_ represents the frozen encoder parameters and Pool(·) denotes the mean-pooling operation over the sequence length, resulting in a static feature vector **v**_*i*_ ∈ ℝ^1024^.

#### 4.2.1. MLP Classification Head

To map high-dimensional semantic representations into clinical categories, static embeddings **v**_*i*_ are processed through a MLP head. The architectural transformation is formalized as:

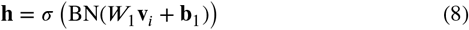

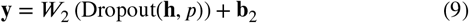

where 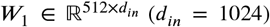 and *W*_2_ ∈ ℝ^*K*×512^ represent the weight matrices, **b** denotes the bias vectors, and *σ* is the activation function of the Rectified Linear Unit (ReLU). During the training phase, the pre-trained transformer parameters Θ remain frozen, serving as a static feature extractor. Consequently, the optimization objective is restricted to the MLP parameter set *ϕ* = {*W*_1_, **b**_1_, *W*_2_, **b**_2_}.The input embedding **v**_*i*_ is derived as:

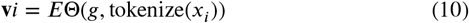

where *E*_Θ_ denotes the frozen INSTRUCTOR-XL encoder, *g* is the task-specific instruction prompt and *x*_*i*_ represents the psychiatric text input. After mean-pooling over the token embeddings, the resulting fixed-length latent representation is

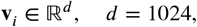

which serves as a static feature vector for the MLP classifier. Importantly, all encoder parameters are frozen during training, preserving pretrained semantic representations, and using the encoder strictly as a feature extractor.

#### 4.2.2. MLP Classification Head

The frozen embeddings **v**_*i*_ are passed to a shallow MLP classifier that predicts one of the psychiatric categories *K* (*K* = 4). The forward pass is defined as

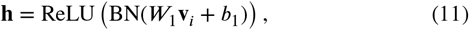

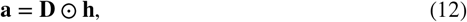

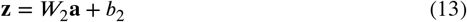

where *W*_1_ ∈ ℝ^512×1024^, *W*_2_ ∈ ℝ^*K*×512^, BN denotes batch normalization and **D** is an element-wise dropout mask with drop probability *p* = 0.4. The output logits **z** are optimized using categorical cross-entropy loss, with soft-max normalization implicitly incorporated into the loss function. Only the MLP parameters *ϕ* = *W*_1_, *b*_1_, *W*_2_, *b*_2_ are updated during training.

#### 4.2.3. Optimization and Training Dynamics

The MLP parameters *ϕ* were optimized using the AdamW optimizer with a learning rate *η* = 1 × 10^−3^. Training was conducted for 10 epochs, minimizing the categorical cross-entropy loss (unweighted):

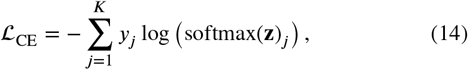

where *y*_*j*_ is the one-hot encoded ground-truth label and **z** are the output logits for each class. The model hyperparameters are reported in Table 4. Model performance was evaluated using accuracy and macro-averaged F1 score.

**Table 4.**
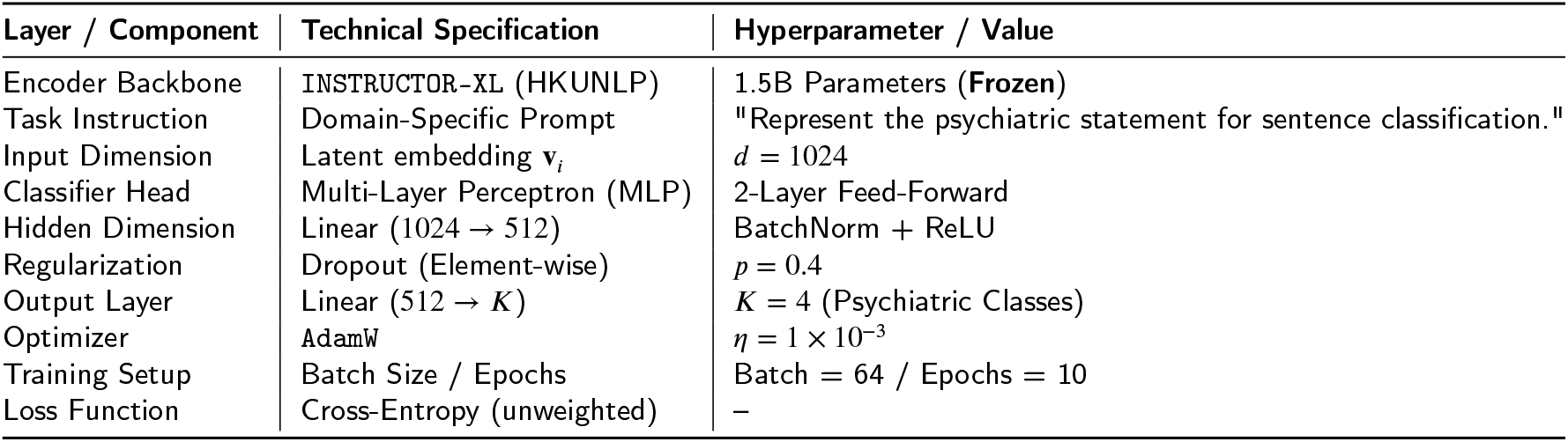
Model B Architecture: Frozen INSTRUCTOR-XL Encoder + MLP Head.

| Layer / Component | Technical Specification | Hyperparameter / Value |
| --- | --- | --- |
| Encoder Backbone | INSTRUCTOR-XL (HKUNLP) | 1.5B Parameters (**Frozen**) |
| Task Instruction | Domain-Specific Prompt | "Represent the psychiatric statement for sentence classification." |
| Input Dimension | Latent embedding $\mathbf{v}_i$ | $d = 1024$ |
| Classifier Head | Multi-Layer Perceptron (MLP) | 2-Layer Feed-Forward |
| Hidden Dimension | Linear ($1024 \rightarrow 512$) | BatchNorm + ReLU |
| Regularization | Dropout (Element-wise) | $p = 0.4$ |
| Output Layer | Linear ($512 \rightarrow K$) | $K = 4$ (Psychiatric Classes) |
| Optimizer | AdamW | $\eta = 1 \times 10^{-3}$ |
| Training Setup | Batch Size / Epochs | Batch = 64 / Epochs = 10 |
| Loss Function | Cross-Entropy (unweighted) | – |

## 5. Results and Performance Analysis

We evaluated two architectures—fine-tuned Bio-ClinicalBERT refined (Model A) and Instructor-XL high-capacity (Model B)—on a clinical corpus (*N* = 151, 228) using train/test/validation k-fold cross-validation operation. Given class imbalance, performance was assessed via Matthews Correlation Coefficient (MCC) and Macro-*F*_1_, emphasizing the models’ ability to discriminate between psychiatric categories.

### 5.1. Model A Performance: The Clinical Specialist Paradigm

Model A (Bio-ClinicalBERT, 110M parameters) achieved 87% validation accuracy and a Macro-*F*_1_ score of 0.782 across the four psychiatric categories, reflecting robust performance despite severe class imbalance (e.g., *suicidal intention n* = 12,783 vs. *schizophrenia n* = 100). Precision for *suicidal intention* reached 1.00, indicating no false-positive predictions, while recall was 0.864, showing that some cases were not detected. The model also demonstrated high recall for *depression* (0.941) and strong performance on *anxiety* (*F*_1_ = 0.826), suggesting effective sensitivity to prevalent psychiatric narratives. Schizophrenia predictions were more limited (*F*_1_ = 0.732), reflecting the challenges imposed by extreme underrepresentation. The confusion matrix indicates that the majority of errors involve misclassifications between clinically overlapping categories, such as depression and suicidal intention. Overall, Model A maintained a balanced discrimination across classes as evidenced by its Macro-F1 and MCC of 0.6752, highlighting that domain-specific fine-tuning improved sensitivity to clinically relevant signals while mitigating the impact of severe class imbalance.

### 5.2. Model B Performance: The Generalist Baseline

Model B employs the 1.5-billion parameter `Instructor-XL` transformer with frozen encoder weights (∇_Θ_ = 0), projecting psychiatric narratives into a fixed 1024 dimensional latent space. This design tests whether large-scale general-purpose pretraining can capture clinically relevant features without domain-specific fine-tuning. As shown in Table 5, Model B achieved 84% validation accuracy, demonstrating that generalist embeddings capture a substantial portion of the task-relevant signal. However, its Matthews correlation coefficient (MCC = 0.6169) is lower than Model A (0.6752), indicating reduced sensitivity to category-specific linguistic markers and clinically salient vocabulary. This is most pronounced for the *depression* category (*F*_1_ = 0.587, AUC = 0.916), where misclassifications occur predominantly with *suicidal intention* (329 cases), reflecting overlapping symptomatology and the limited ability of a frozen generalist model to disambiguate subtle phenotypic distinctions. Performance on other categories, such as *schizophrenia* (*F*_1_ = 0.798) and *anxiety* (*F*_1_ = 0.762), is comparatively robust, suggesting that broad semantic priors are sufficient for less frequent or more lexically distinct categories. Overall, Model B demonstrates competitive but lower discriminative performance relative to Model A, highlighting the importance of domain-specific fine-tuning for capturing nuanced clinical semantics in psychiatric text.

**Table 5.** Validation Performance Metrics Comparison: Bio-ClinicalBERT (Model A) vs. Instructor (Model B) for Multi-Class Psychiatric Categories Classification.

| Clinical Psychiatric Categories | Model A: Bio-ClinicalBERT |  |  |  | Model B: Instructor |  |  |  |
| --- | --- | --- | --- | --- | --- | --- | --- | --- |
|  | Prec. | Rec. | F1 | Supp. | Prec. | Rec. | F1 | Supp. |
| Anxiety | 0.772 | 0.893 | 0.826 | 572 | 0.682 | 0.862 | 0.762 | 572 |
| Depression | 0.483 | 0.941 | 0.637 | 1668 | 0.483 | 0.763 | 0.587 | 1668 |
| Schizophrenia | 0.591 | 0.972 | 0.732 | 100 | 0.694 | 0.952 | 0.798 | 100 |
| Suicidal | 1.00 | 0.864 | 0.922 | 12783 | 0.971 | 0.881 | 0.924 | 12783 |
| **Overall Accuracy** |  | 0.87 |  | 15123 |  | 0.84 |  | 15123 |
| **Macro Average** | 0.711 | 0.922 | 0.782 | 15123 | 0.712 | 0.863 | 0.773 | 15123 |
| **Weighted Average** | 0.932 | 0.871 | 0.891 | 15123 | 0.904 | 0.872 | 0.882 | 15123 |

Note: Support refers to the validation sample size ( $n$ ). Model A is Bio-ClinicalBERT (110M parameters); Model B is Instructor (frozen generalist, 1.5B parameters). Precision, Recall, and F1-Score are reported as decimals.

### 5.3. Comparative Statistical Analysis

The performance of both models across psychiatric categories was evaluated on the validation set (Table 2), with detailed results summarized in Table 6. Model A exhibited higher *F*_1_-scores for *anxiety* (*F*_1_ gain = 0.064) and *depression* (*F*_1_ gain = 0.050), suggesting that domain-specific fine-tuning may improve classification for these relatively well-represented categories. In contrast, Model B (`Instructor-XL`) achieved the highest *F*_1_ for *schizophrenia* (*F*_1_ = 0.798). Given the small sample size for this class (*n* = 100), this difference should be interpreted cautiously and may reflect the contribution of large-scale pretraining to performance on underrepresented categories. Overall, these results indicate that domain-specific fine-tuning and generalist pretraining confer complementary advantages in capturing heterogeneous linguistic patterns across psychiatric categories.

**Table 6.** Class-Specific Performance Comparison: Model A vs. Model B.

| Phenotype | Model A $F_1$ | Model B $F_1$ | Winner | Margin |
| --- | --- | --- | --- | --- |
| Anxiety | 0.826 | 0.762 | Model A | 0.064 |
| Depression | 0.637 | 0.587 | Model A | 0.050 |
| Schizophrenia | 0.732 | 0.798 | Model B | 0.066 |
| Suicidal | 0.922 | 0.924 | Model B | 0.002 |

Note: Scores based on $F_1$ decimals from Table 5.

### 5.4. XAI Evaluation: Specialist Gaze vs. Semantic Diffusion

To interpret the differences in model performance (MCC = 0.6752 for Model A vs. 0.6169 for Model B), we performed a feature attribution analysis using gradient-based saliency maps. This analysis provides insight into how each model assigns importance to specific lexical and semantic features within psychiatric texts. Preliminary qualitative inspection indicates that the models emphasize different aspects of the input: one model predominantly highlights domain-specific terms, whereas the other captures broader semantic patterns across the text. These observations inform downstream interpretability and guide further investigation of model decision-making strategies.

#### 5.4.1. Salience Map Analysis

We extracted token-level attention weights from the last hidden layer of two transformer-based models—Model A (specialist) and Model B (generalist)—for representative schizophrenia and depression text samples (Fig. 5c). Attention weights were normalized to the [0,1], providing an approximate indication of the model’s internal focus across tokens. Qualitative inspection reveals that Model A tends to assign higher attention to tokens associated with psychiatric symptom markers, reflecting structured co-occurrence patterns relevant to diagnosis. In contrast, Model B exhibits more diffuse attention distributions, with less alignment to clinically relevant tokens. Attention values were visualized using color-coded bar plots overlaid with ground-truth and predicted labels, enabling comparison between model focus and known symptom-relevant terms. These analyses provide a qualitative assessment of model interpretability and suggest systematic differences in how specialist and generalist models encode token-level information related to psychiatric features. The results indicate an architectural split:

- **Model-A:** Achieves strong performance in *Anxiety* and *depression*. Fine-tuning enables the model to resolve subtle semantic overlaps typically observed in narratives about affective disorders.
- **Model-B:** Exhibits greater robustness in the *schizophrenia* category. The extensive 1.5B-parameter pretraining provides stable latent representations for rare classes with limited training examples.

## 6. Discussion

The divergent performance trajectories of the two architectures provide significant insights into the mechanisms of semantic feature fusion within psychiatric discourse. Attention-based saliency maps (Fig. 5c) demonstrate that both models prioritize tokens associated with specific psychiatric semiology. This suggests that the latent representations do not simply capture spurious lexical correlations but are aligned with the structured manifold of psychiatric language. The substantial class imbalance (*n*_suicide_ = 12, 783 vs. *n*_schizophrenia_ = 100) requires a shift from global accuracy toward more robust diagnostic contingency metrics. In this context, the MCC serves as a measure of the information-theoretic reliability of the classifiers (38). The observed superiority of model A in underrepresented categories suggests that task-specific fine-tuning facilitates a more efficient collapse of the high-dimensional clinical space into discriminative clusters. By bypassing predefined lexicons (e.g., LIWC), Bio-ClinicalBERT performs an end-to-end fusion of raw clinical text and domain-specific priors, encoding associations between symptoms and labels that are often lost in discrete, lexicon-based mapping approaches (8). The results highlight a trade-off between local domain adaptation (Model A) and global semantic robustness (Model B): Phenotypic Density: Model A’s higher F1-scores for anxiety and depression indicate that fine-tuning is superior for phenotypes characterized by high “linguistic density”—where symptoms are expressed through subtle variations in common language. Here, the model must refine its attention weights to distinguish between colloquial distress and clinical pathology. In contrast, Model B’s resilience in the schizophrenia category—despite the data sparsity—demonstrates the benefit of cross-task knowledge fusion. The INSTRUCTOR-XL backbone utilizes instructional priors to stabilize the latent representation of rare, distinctive linguistic markers. This indicates that for low-resource psychiatric categories, the fusion of large-scale instructional pretraining and static feature extraction provides a more stable substrate for classification, mitigating the risk of catastrophic forgetting or overfitting inherent in fine-tuning small samples (39). These findings suggest a hierarchical approach to psychiatric information fusion. While fine-tuned models better capture overlapping symptom clusters in high-density data, instruction-tuned frozen encoders provide stable representations that preserve the broad semantic structure necessary to identify low-frequency, high-salience diagnostic markers. Combining domain-specific fine-tuning with generalist pretrained encoders offers a framework for building more reliable diagnostic models under realistic clinical data conditions.

## 7. Conclusion

Our results reveal a fundamental trade-off in psychiatric NLP: while domain-specific fine-tuning (Model A) resolves the subtle linguistic nuances separating anxiety from depression, a frozen generalist encoder (Model B) provides the structural stability necessary to prevent overfitting in low-resource conditions like schizophrenia. This dual-model framework demonstrates that patient-generated narratives, whether from clinical interviews or handwritten reports, encode high-dimensional cognitive and affective phenotypes that are often lost in categorical scaling. By modeling the semantic structure of psychiatric interviews and written patient narratives, we move beyond simple binary classification toward fine-grained longitudinal monitoring of clinical change. This is particularly critical for capturing cross-modal dissociations in treatment-resistant populations, where subjective narrative distress may persist despite the stabilization of conventional clinical markers. Moving forward, our work at McLean Hospital will integrate these linguistic signatures with objective behavioral and neurophysiological data. This multimodal approach aims to transition psychiatric evaluation from reactive, snapshot-based assessments to a proactive, data-driven framework capable of tracking the evolution of complex mental health trajectories in real-world contexts.

## Data Availability

All data produced in the present study are available upon reasonable request to the authors. All data produced in the present work are contained in the manuscript.

## Foundings

This work was financially supported by Dr. Giuseppe Varone, who provided funding for computational resources, data management, and analysis infrastructure. Dr. Joshua C. Brown and Dr. Varone also acknowledge support from the Defense Advanced Research Projects Agency (DARPA)-funded project BBRF #GR0402118.

## CRediT authorship contribution statement

**Giuseppe Varone:** Conceptualization, Methodology, Software Development, Data Analysis, Writing – Original Draft, Funding Acquisition. **Poornima Kumar:** Writing - Original draft preparation, Supervision. **Joshua C. Brown:** Writing - Original draft preparation, Supervision. **Wadii Boulila:** Writing - Original draft preparation, Methodology, Supervision.

---

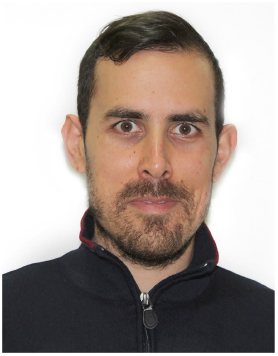

Giuseppe Varone is a senior postdoctoral research fellow at McLean Hospital and Harvard Medical School. His research focuses on pharmaco-TMS-EEG, synaptic mechanisms, and deep learning applied to psychiatric disorders. He received a B.S. in Computer Science, M.S. degrees in Biomedical and Electronics Engineering, and a Ph.D. in Biomarkers of Chronic and Complex Diseases. His work includes optimization of TMS-EEG acquisition systems, brain stimulation hardware–software integration, biofeedback, closed-loop brain stimulation, and computational psychiatry. He is a Senior Member of IEEE.

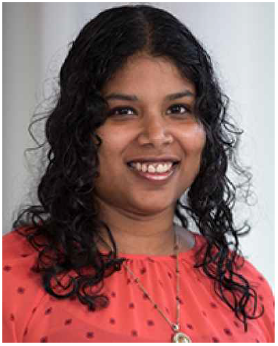

Poornima Anandh Kumar, PhD, is an assistant professor of psychiatry at Harvard Medical School and an assistant neuroscientist at McLean Hospital. Her research focuses on investigating the mechanisms through which humans learn and process reinforcements (both rewards and punishments) and how these processes might contribute to psychiatric disorders, particularly depression. She utilizes computational modeling and multimodal imaging techniques including fMRI, EEG, and MRS to better understand the clinical and neurobiological correlates of reinforcement learning. Core features of depression are anhedonia (loss of pleasure) and negative bias (excessive attention and memory for negative events), which might be due to abnormal reinforcement learning.

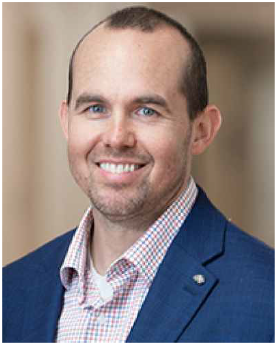

Joshua C. Brown, MD, PhD, is a psychiatrist, neurologist, and neuroscientist. He is medical director of the Transcranial Magnetic Stimulation (TMS) Service and director of TMS research in the Division of Depression and Anxiety Disorders at McLean Hospital. Dr. Brown is also director of the Brain Stimulation Mechanisms Laboratory at McLean and an assistant professor of Psychiatry at Harvard Medical School. He is internationally recognized for his work on synaptic mechanisms of TMS, pharmacologic augmentation of TMS, and TMS parameter selection. He is the founding editor-in-chief of the journal Transcranial Magnetic Stimulation and president-elect of the Clinical TMS Society.

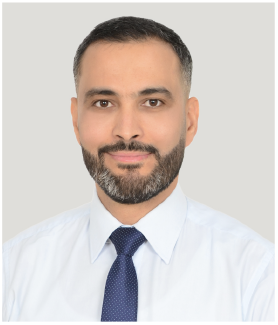

Wadii Boulila received his B.Sc. degree (First-Class Honours with Distinction) in Computer Science from the Aviation School of Borj El Amri in 2005, his M.Sc. degree in Computer Science from the National School of Computer Science (ENSI) at the University of Manouba, Tunisia, in 2007, and his Ph.D. in Computer Science, University of Rennes 1, France, in 2012. He is currently a Professor of Computer Science and the director of the RIOTU Laboratory at Prince Sultan University, Saudi Arabia. His research interests include data science, computer vision, big data analytics, deep learning, cyber-security, artificial intelligence, and uncertainty modeling. Prof. Boulila is a Senior Member of IEEE, Member of ACM and AAAI, and a Senior Fellow of the Higher Education Academy (SFHEA), U.K.

